# Effect of an educational intervention on knowledge, attitude, and self-efficacy of child sexual abuse prevention among parents in Ogun State, Nigeria

**DOI:** 10.1101/2025.08.07.25333253

**Authors:** Comfort Adebisi Ogunleye, Joel Aluko, Rebecca Oluwatosin Oyedele, Temitope Elizabeth Opaleye, Ezekiel Olusola Omitogun, Grace Orunmuyiwa, Ayodeji Olubunmi Ogunmuyiwa, Blessed Ayobola Ojumu, Oluwadamilare Akingbade

**Affiliations:** Department of Nursing Sciences, Babcock University, Ilishan, Ogun State, Nigeria; Ogun State College of Nursing Sciences, Ilaro, Ogun State, Nigeria; Department of Nursing, University of Ilorin, Kwara State, Nigeria; Institute of Nursing Research, Osogbo, Osun State, Nigeria; School of Psychiatric Nursing, Neuropsychiatric Hospital Aro Abeokuta, Ogun State, Nigeria; Department of Nursing Science, Olabisi Onabanjo University, Sagamu Campus, Ogun State, Nigeria; Department of Nursing, Lagos State University Teaching Hospital, Lagos State, Nigeria; Faculty of Nursing, University of Alberta, Edmonton, Canada

## Abstract

**Background:** Parents’ efforts are essential in the prevention of Child Sexual Abuse, as studies consistently underscore their critical roles in protecting and providing a safe place for their children. This study highlights the pivotal role of parents in preventing child sexual abuse, as research consistently emphasizes their responsibility in safeguarding their children and creating a secure environment. However, despite their critical role, studies indicate that parents often lack sufficient knowledge, exhibit poor attitudes, and demonstrate low self-efficacy regarding child sexual abuse prevention, contributing to higher child sexual abuse incidence. To address this gap, the study investigated the effect of an educational intervention on parents’ knowledge, attitudes, and self-efficacy related to child sexual abuse prevention in Ogun State, Nigeria.

**Method:** A quasi-experimental design was conducted in two selected Junior Secondary Schools in Yewa South and Abeokuta South Local Government Areas, Ogun State, Nigeria. It involved an intervention group and a control group with a sample size of 249 selected by a systematic random sample. Data were collected using a validated questionnaire. Assessments were conducted at baseline [P0], four weeks post-intervention [P1], and twelve weeks post-intervention [P2]. Data were analyzed using descriptive and inferential statistics at a 0.05 significance level.

**Results:** Results revealed significant improvements [p < 0.05] in knowledge, attitudes, and self-efficacy were observed in the intervention group post-intervention. The Cohen’s D effect size for knowledge, attitude, and self-efficacy of child sexual abuse prevention were 0.27, 0.54, and 0.57, respectively.

**Conclusion:** The study demonstrated the efficacy of the educational intervention in improving parents’ knowledge, attitudes, and self-efficacy regarding child sexual abuse prevention. It is recommended that educational interventions be widely implemented to raise awareness and empower parents to effectively prevent child sexual abuse in their communities.

## INTRODUCTION

Child Sexual Abuse (CSA) remains a pervasive global issue with profound immediate and long-term consequences for victims, necessitating a multifaceted and comprehensive prevention strategy. Parents, as primary caregivers, play a critical role in safeguarding children from Child Sexual Abuse (CSA), particularly during adolescence when susceptibility to abuse increases due to physical, emotional, and social developmental changes [1]. However, research indicates that many parents lack the necessary knowledge, exhibit negative attitudes, and demonstrate low self-efficacy in preventing Child Sexual Abuse (CSA), creating a significant gap in their ability to protect their children effectively [2–4]. Structured educational interventions have been proposed to empower parents with the requisite skills, foster positive attitudes, and enhance their self-efficacy, thereby creating safer environments for children and reducing CSA incidence [5].

Globally, CSA prevalence remains alarmingly high, with 7.9% of males and 19.7% of females reporting experiences of abuse [6]. Africa reports some of the highest rates, ranging from 2.1% to 68%, with Nigeria experiencing significant prevalence rates of 15% to 38% across various regions [7, 8]. In Ogun State, Nigeria, CSA prevalence is estimated at 33.57%, with 111 cases reported in Ilaro and 71 in Abeokuta in 2021 alone [9]. Despite these staggering statistics, CSA remains underreported due to cultural taboos, fear of stigma, and limited awareness, exacerbating its devastating impacts on victims. These include physical injuries, sexually transmitted infections, unwanted pregnancies, psychological trauma, and long-term health issues such as depression, substance abuse, and chronic diseases [10].

Parents’ knowledge, attitudes, and self-efficacy are critical determinants in CSA prevention. Adequate knowledge enables parents to recognize warning signs, intervene promptly, and establish protective measures, while positive attitudes foster open communication with children about sensitive topics [5]. Self-efficacy, the belief in one’s ability to take effective action, empowers parents to advocate for their children, report suspicions, and seek professional help when necessary [11]. However, many parents rely on inadequate sources of information, such as social media, and face challenges in addressing children’s questions about sexual health, further highlighting the need for structured educational interventions [12, 13].

The Social Cognitive Theory [14] provides a theoretical framework for understanding how educational interventions can enhance parents’ knowledge, attitudes, and self-efficacy. This theory posits that learning occurs through reciprocal interactions between personal, behavioral, and environmental factors, suggesting that parents can develop confidence and skills through observation, reinforcement, and feedback [14]. By equipping parents with comprehensive sexual education, including biological, cultural, and psychological aspects, interventions can address cognitive, emotional, and behavioral dimensions, fostering a balanced understanding of sexuality and promoting protective behaviors [12, 15].

Despite the critical role of parents in CSA prevention, limited studies have explored the effectiveness of educational interventions in Nigeria. Knowledgeable parents can implement prevention programs to educate children about what is the appropriate behaviour, and thus reduce the likelihood of them becoming victims. Also, well-informed parents with the right attitude can advocate for stronger child protection policies and resources within the community and educational institutions. They can also provide support for victims and their families. Improvement in these areas can lead to better prevention practices and better support for affected children. Hence, this study assessed the effectiveness of a structured educational intervention on the knowledge, attitude, and self-efficacy of parents in preventing CSA.

## METHODS

### Study design and setting

This study adopted a quasi-experimental design with a pre-intervention and post-intervention study conducted on two groups (intervention and control group). The study was conducted in Ogun State, Nigeria. The target population was parents of children in two selected secondary schools, Yewa College, Ilaro, and Abeokuta Grammar School, Abeokuta. These schools are located in areas with the highest reported cases of CSA in the State [9].

### Sample and Sampling Technique

The sample size for this study was determined using the formula for calculating the difference in means [16]. A sample size of 249 parents was calculated to ensure statistical precision. A multi-stage sampling approach was employed. In Stage 1, purposive sampling was used to select two senatorial districts out of the three in Ogun State, based on the historical high incidence of Child Sexual Abuse (CSA) cases. Ogun Central and Ogun West senatorial districts were purposively selected due to their high reported cases of CSA (Ogun State Judiciary, 2022). In Stage 2, purposive sampling was again used to select one local government area (LGA) from each senatorial district based on the highest reported CSA cases. The selected LGAs were Yewa South and Abeokuta South.

In Stage 3, the purposive sampling technique was used to select the ward with the highest reported CSA cases within each LGA. The selected wards were Ilaro I and Ijaiye/Idi-aba. In Stage 4, purposive sampling was used to select one public secondary school from each of the two wards based on the highest student populations in the respective LGAs. The two schools selected were Yewa College Ilaro and Abeokuta Grammar School Idi-aba. In Stage 5, paper balloting was conducted to randomly assign the selected schools to the experimental and control groups. Yewa College Ilaro was assigned as the experimental group, while Abeokuta Grammar School Idi-aba was designated as the control group. In Stage 6, a proportionate sampling technique was used to select students from the three junior secondary classes (JSS1, JSS2, and JSS3) in each school. In Stage 7, a systematic sampling technique was employed, using the enrollment list as the sampling frame. Every nth (20th) student in the class was selected and approached for participation. Only students whose parents provided positive responses and informed consent, along with approval for participation, were included in the study.

Selected parents were then randomly assigned to either the Intervention Group (IG) or the Control Group (CG). There were 126 participants in the Intervention Group (IG) and 123 participants in the Control Group (CG) respectively. The recruitment of participants started on 15^th^ August, 2024, and ended on 4^th^ October, 2024. The inclusion criteria are parents, guardians, caregivers and foster parents who signed informed consent, above 18years, who can communicate fluently in English or Yoruba language, who are willing to participate and complete the pre and post assessments. Parents who had gone through any previous training on CSA prevention, parents who were too sick to answer questions and those who had plans to relocate during the period of the study were excluded from the study.

### Instrumentation and data collection

A structured questionnaire was administered to the selected participants to collect data. The Turkish adaptation of the Child Sexual Abuse Knowledge/Attitude Scale Questionnaire (CSAKAS) was utilized to evaluate parents’ knowledge and attitudes regarding child sexual abuse prevention [17]. Additionally, the Farell and Walsh Parental Efficacy Questionnaire was employed to measure parents’ self-efficacy in the prevention of CSA [5, 18]. These instruments were administered at baseline before intervention (P0), four weeks post-intervention (P1), and twelve weeks post-intervention (P2). The combined tool comprised a total of 55 question items under 4 sections (Sections A-D). Section A assessed the socio-demographic data of the participants, Section B assessed the participant’s knowledge of CSA with dichotomous questions of True/False. Section C and D assessed the attitude and self-efficacy of parents in CSA prevention respectively, based on a 4-level Likert scale. At the pre-intervention phase, participants were grouped by their level of literacy, questionnaires were administered and filled with the help of the research assistants who had been trained prior time. The questions with the options were read for the illiterates, they were allowed to pick their choice of responses and same documented. The questionnaire (English or Yoruba version) was used for data collection in both experimental and control groups. Following the intervention, post-test questionnaire was administered after the fourth- and twelfth-week post-intervention using the same questionnaire as in the baseline data collection.

### Intervention

An educational module was prepared as an intervention package for this study. There were consultations with experts in the field of education and child protection agencies. Also, documentary evidence from Child Sexual Abuse Prevention guide developed by United Nations Children’s Fund [19] and Centre for Disease Control CSA prevention policy checklist was used to develop the intervention package [20]. The intervention was exclusively administered to the intervention group and comprised four weekly sessions, each lasting 90 minutes. The sessions were designed to be interactive and participatory, incorporating lectures, question-and-answer segments, role-playing activities, and group discussions. These methods encouraged parents to explore, discuss, and practice effective parenting strategies for preventing CSA. Through group work, parents had the opportunity to listen to others, share similar parenting experiences, and adapt approaches to their unique family contexts. This collaborative environment provided confirmation and support, enabling parents to refine their practices and build confidence in their ability to protect their children. The content of the education was structured around specific behavioural and educational objectives that involved improvement in knowledge and awareness of preventing CSA, right attitude to child protection/sexual abuse prevention and self-efficacy in preventing child sexual abuse. Topics related to sexual abuse such as types of sexual abuse, frequency, effects, identification of possible indicators related to sexual abuse, characteristics of both the victim and perpetrator, high-risk situations, false beliefs, and appropriate action in the face of possible sexual abuse, available community resources were discussed. The participants were given opportunity to ask questions for clarifications. Information Education Communication (IEC) leaflet was distributed to the participants to take home to enhance their memory on what has been taught on prevention of CSA.

### Statistical Analysis

The collected data were analyzed using the International Business Machine-Statistical Package for Social Sciences (IBM-SPSS, Armonk, NY, USA) version 23. The data were initially coded into the software and subsequently cleaned to ensure accuracy. Both descriptive and inferential statistical methods were employed for data analysis. Descriptive statistics, including frequency distribution tables, percentages, means, and standard deviations, were used to summarize the sociodemographic characteristics of the participants and address the research questions. For inferential analysis, paired t-tests were conducted to compare variables within and between groups, while Pearson Product-Moment Correlation (PPMC) was used to test the hypotheses. The effect size was measured using Cohen’s D. All hypotheses were tested at a 0.05 level of significance to determine statistical significance.

### Ethical Considerations

An ethical clearance and approval were obtained from Babcock University Research and Ethical Committee (BURHEC) with reference number BUHREC 808/23 based on institutional requirements and also, from the Ogun State Health Research Committee (OGHREC) with reference number OGHREC/467/205. Verbal and written consent were obtained from all the study participants. Their anonymity and respect for autonomy were also assured.

## RESULT

A total of 249 parents participated in the quasi-experimental component of the study, comprising of 126 participants in the Intervention Group (IG) and 123 participants in the Control Group (CG). The response rate was 97.7% and 95.3% for the intervention and control group, respectively. Table 1 shows the demographic characteristics of the participants. Females constitute the highest proportion in both the intervention and control groups. Majority (85%) of participants in the intervention group and 69% of the participants in the control group were parents.

**Table 1.** Socio-demographic characteristics of the respondents in the intervention and control group.

| **Variables** | **Intervention Group (IG)**<br>**N= 126**<br>**N (%)** | **Control Group (CG)**<br>**N=123**<br>**N (%)** |
| --- | --- | --- |
| **Age(years)** |  |  |
| 15-24 | 3(3.3) | 18(14.6) |
| 25-34 | 17(13.5) | 27(22.0) |
| 35-44 | 66(52.4) | 52(42.3) |
| 45-54 | 32(25.4) | 19(15.4) |
| 55-64 | 8(6.4) | 6(4.9) |
| 65-74 | 0(0) | 1(0.8) |
| **Mean±SD** | **41.97±7.47** | **37.28±10.73** |
| **Sex** |  |  |
| Male | 32(24.8) | 31(25.2) |
| Female | 94(74.6) | 92(74.8) |
| **No of Children** |  |  |
| 0-3 | 65(51.6) | 95(77.2) |
| 4-8 | 61(48.4) | 28(22.8) |
| **Relationship to child** |  |  |
| Parent | 107(84.9) | 85(69.1) |
| Guardians | 15(11.9) | 30(24.4) |
| Others | 4(3.2) | 8(6.5) |
| **Ethnicity** |  |  |
| Yoruba | 97(77.0) | 81(65.9) |
| Igbo | 16(12.7) | 30(24.3) |
| Hausa | 9(7.1) | 4(3.3) |
| Others | 4(3.2) | 8(6.5) |
| **Level of Education** |  |  |
| No formal | 12(9.5) | 4(3.3) |
| Primary | 27(21.4) | 16(13.0) |
| Secondary | 62(49.2) | 23(18.7) |
| Post-secondary | 3(2.4) | 25(20.3) |
| University | 22(17.5) | 55(44.7) |
| **Occupation** |  |  |
| Currently fully employed | 48(38.1) | 66(53.7) |
| Employed part-time | 43(34.1) | 38(30.9) |
| Currently unemployed | 29(23.0) | 11(8.9) |
| Retired | 6(4.8) | 8(6.5) |
| **Nature of Occupation** |  |  |
| Civil servant | 15(15.1) | 30(24.4) |
| Business /self-employed | 89(70.6) | 60(50.4) |
| Professional | 14(11.1) | 28(22.8) |
| House wife | 1(0.8) | 1(0.8) |
| Retired | 3(2.4) | 2(1.6) |

The result of the participants’ level of knowledge, attitude and self-efficacy on CSA at pre-intervention (P0), and evaluation at 4^th^ (P1) and 12^th^ (P2) week post-intervention in the Control and Intervention Group were analyzed on Table 2. At Pre-Intervention (P0), the mean ± SD score for the participants level of knowledge regarding child sexual abuse in the control and intervention group was 13.28 ± 2.39 and 11.86 ± 2.58 respectively. At the 4th week post intervention (P1), the intervention group had an increase in the mean knowledge score from 11.86 ±2.58 at pre-intervention to 17.70 ±1.34. While at 12^th^ week post-intervention (P2), the mean knowledge score of the intervention group was 17.33 ± 1.43, which was slightly lower than the mean knowledge gain experienced by the participants at the 4th week post intervention (17.70 ± 1.34). This reduction in knowledge was insignificant (*p* = 0.05) with minimal effect size. At the control group, there was also a slight increase in the mean score at P1, which was insignificant (*p*= 0.27) and was also followed by a reduction at P2.

**Table 2:**
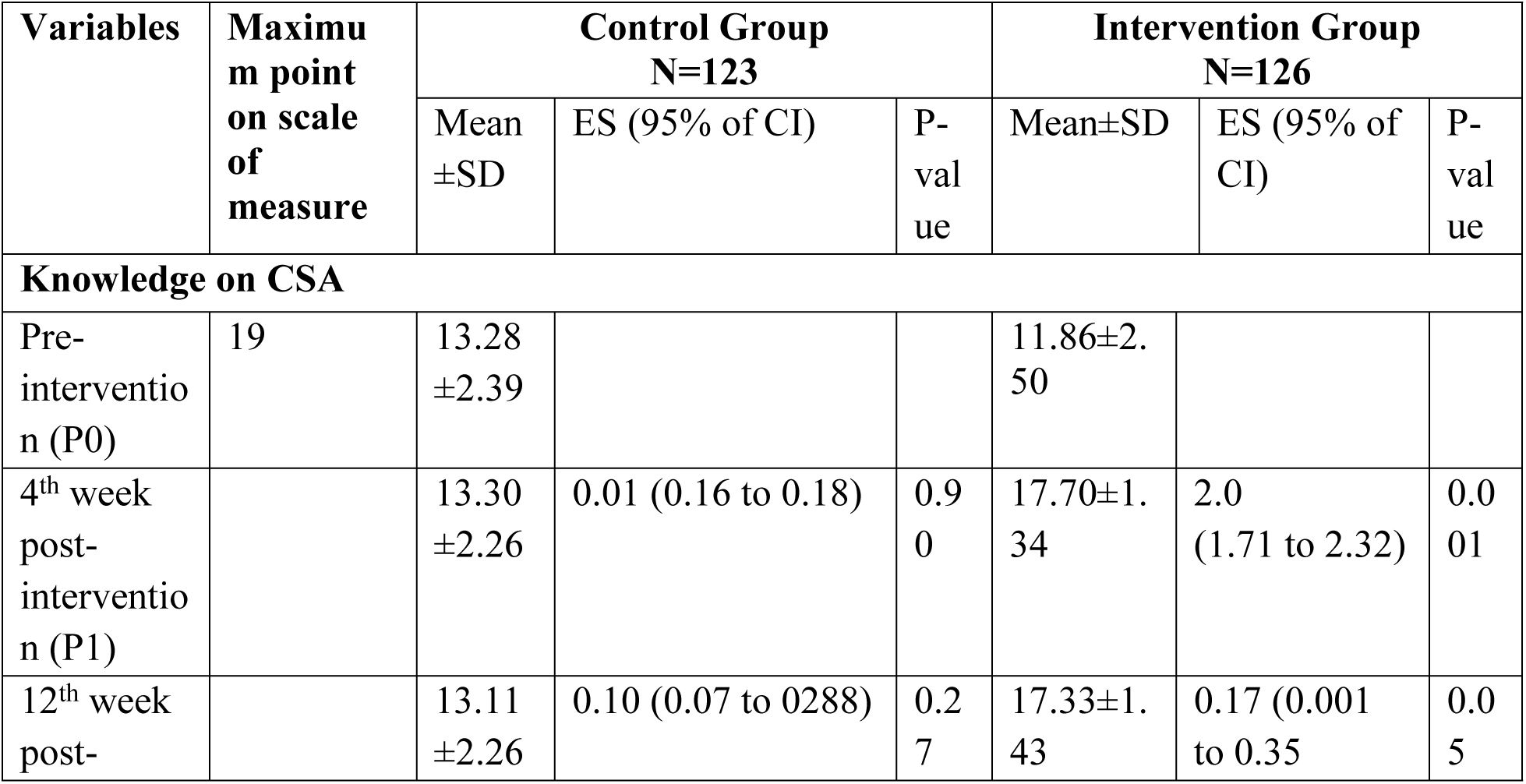
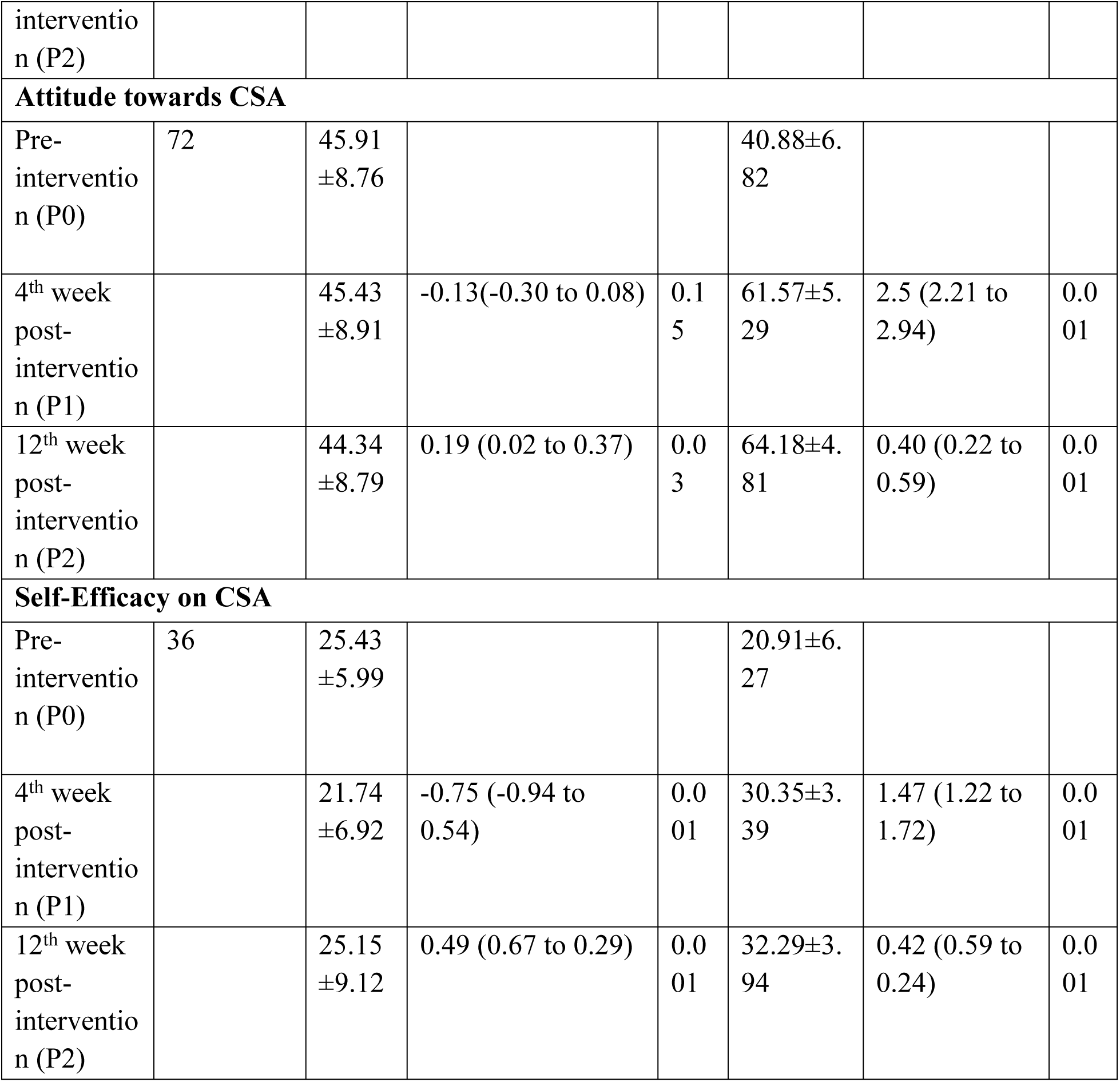
Categorization of the Participants Level of Knowledge, attitude and self-efficacy on Child Sexual Abuse at Pre-Intervention, and evaluation at 4^th^ and 12^th^ week post-intervention in the Control and Intervention Group.

The participant’s attitude towards CSA at P1 showed that few of the participants in the control group had negative attitude towards CSA while less than half of the participants in the intervention group had negative attitude towards CSA. At P1, all (100%) the participants in the intervention group had positive attitude towards CSA prevention. The intervention group had an increase in their attitudinal mean score from 40.88 ±6.82 at P1 to 61.57 ±5.29 at P2, while at the control group, there was a slight decrease from 45.91±8.76 to 45.43±8.91. The intervention had an effect size of 2.5 (95% CI: 2.21 to 2.94) on the intervention group, which showed a highly significant (*p* = 0.001) change in the participants attitude towards CSA prevention. At P2, the participants in the intervention group continued to experience an increase in their attitudinal mean score towards CSA prevention from 61.57 ± 5.29 at P1 to 64.18 ± 4.81 at P2. This change in attitude towards CSA prevention experienced by the intervention group was significant (*p* = 0.001) with an effect size of 0.40 (95% CI: 0.22 to 0.59) and there was a continuous decrease experienced by the control group.

The control group had a mean efficacy score of 25.43 ± 5.99 and the intervention group had 20.91 ± 6.27 at P0. At P1, the control group had a reduction to 21.74±6.92, which later got increased to 25.15±9.12 at P2. The intervention group had an increase in self-efficacy mean score from 20.91 ±6.27 at pre-intervention to 30.35 ±3.39. The health education intervention had an effect size of 1.47(95% CI: 1.22 to 1.72) which showed a highly significant (*p* = 0.001) change in the participants self-efficacy of CSA prevention. At 12^th^ week post-intervention (P2), the participants in the intervention group continued to experience an increase in self-efficacy mean score of CSA prevention from 30.35 ± 3.39 at P1 to 32.29 ± 3.94 at P2. This increased in self-efficacy of CSA prevention experienced by the intervention group was significant (*p* = 0.001) with an effect size of 0.42(95% CI: 0.59 to 0.24).

### Test of Hypotheses

Nine hypotheses were tested for this study to determine which of the outcome variables had statistically significant difference at the 4th week post-intervention and 12^th^ week follow-up. Table 3 shows the result of a two-tailed independent sample t-test for statistical significance. There was significant difference between the mean knowledge score, mean attitudinal score and mean self-efficacy score of intervention group and control group at post-intervention, P1 and P2. This implies that the intervention improved the participants’ knowledge, attitude and self-efficacy of CSA prevention.

**Table 3:**
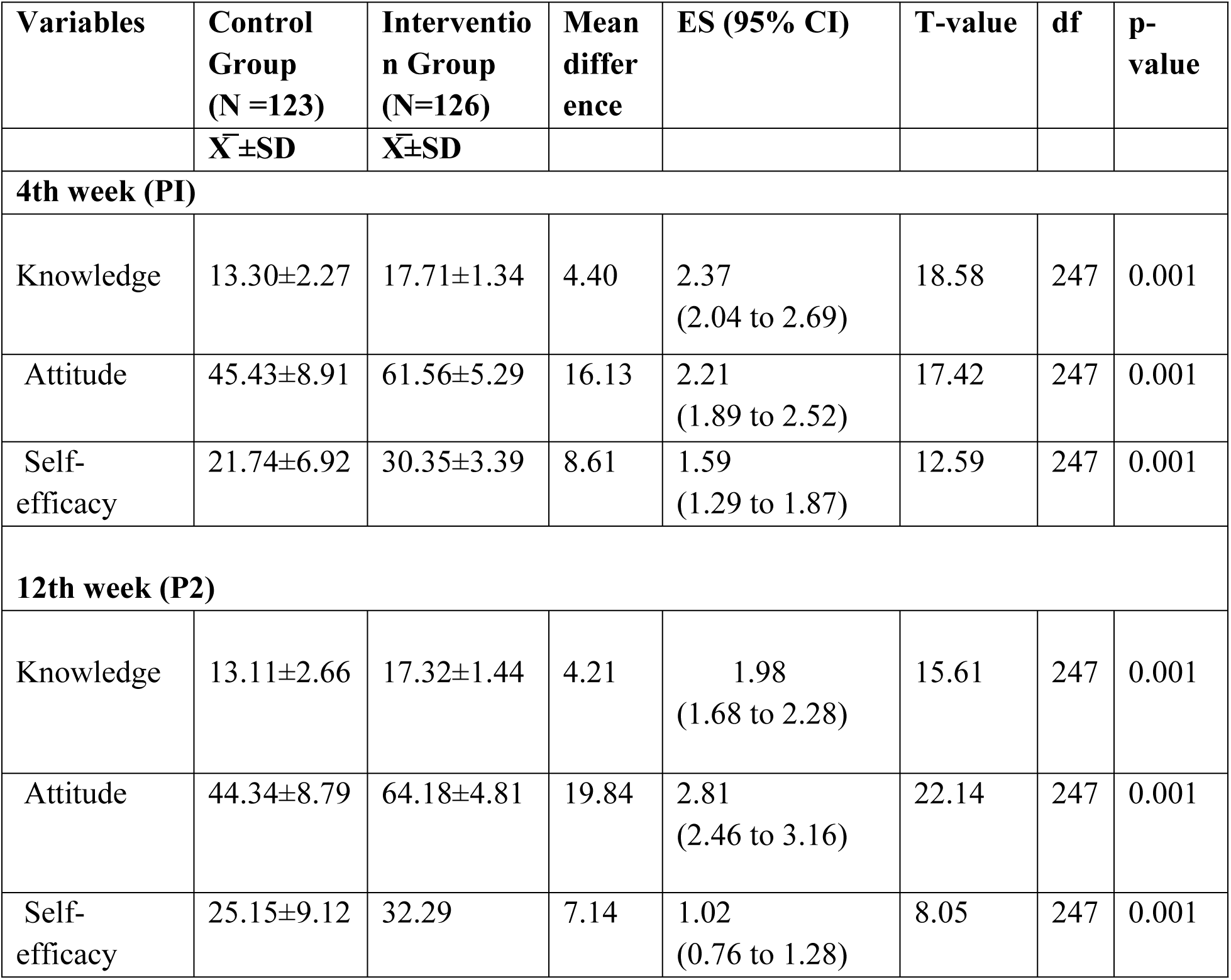
Independent t-test analysis on outcome variables between the control group and the intervention group at post-intervention (P1 and P2)

As shown in Table 4, a paired t-test was conducted to determine if the total mean difference observed in the participants’ knowledge due to the intervention programs is statistically significant. The intervention had a significant effect on the participants’ knowledge, attitude, and self-efficacy regarding CSA prevention in the intervention group. In contrast, the control group showed no statistically significant change in knowledge, a statistically significant decrease in attitude, and no statistically significant change in self-efficacy.

**Table 4:** Paired Sample T-test analysis of variables Regarding Child Sexual Abuse Prevention between Intervention and Control group Pre-intervention and 12^th^ week Post-Intervention.

| Variables | Groups | Mean Diff | S.D | S.E | df | T | *ES(95%CI) | p-value |
| --- | --- | --- | --- | --- | --- | --- | --- | --- |
| **Knowledge regarding CSA** | Intervention | 5.47 | 2.92 | 0.26 | 125 | 20.96 | -1.86(-2.16 to -1.58) | 0.001 |
|  | Control | 0.05 | 3.76 | 0.47 | 122 | 0.98 | 0.09(-0.08 to 0.266) | 0.33 |
| **Attitude towards CSA** | Intervention | 23.30 | 8.79 | 0.78 | 125 | 20.97 | -2.65(-3.01 to -2.27) | 0.001 |
|  | Control | 0.05 | 3.76 | 0.47 | 122 | 2.70 | 0.25(0.07 to 0.43) | 0.006 |
| **Self-efficacy of CSA Prevention** | Intervention | 11.37 | 6.89 | 0.61 | 125 | 18.52 | -1.65(-1.91 to -1.34) | 0.001 |
|  | Control | 0.05 | 3.76 | 0.47 | 122 | 0.44 | 0.04(-0.14 to 0.53) | 0.66 |

## Discussion

This study examined the effect of educational intervention on the knowledge, attitude and self-efficacy on CSA prevention among parents. At baseline, the majority of participants in both the control and intervention groups demonstrated fair knowledge of CSA prevention. This is similar to previous findings by Salloum et al. [21] which reported that most parents were knowledgeable about CSA, viewed CSA prevention as their responsibility, and had talked with their children about CSA. This is further corroborated by Mlekwa et al. [22] who also showed that the majority of respondents had high knowledge regarding prevention of CSA. However, this contrasts with studies that found lower knowledge levels, potentially due to differences in access to information and technological advancements [23, 24].

Following the intervention, knowledge levels in the intervention group improved significantly, majority of participants demonstrating good knowledge at 4 and 12 weeks. This aligns with previous studies which reported similar increase in knowledge following educational interventions [1, 5, 25]. Ferragut et al. [26] further supported these findings, showing a large effect size (dMR = −0.96) for CSA prevention programs in improving knowledge. Interventions in the form of educational interventions have proven to be an effective approach to improving the knowledge of parents about CSA prevention, as the findings of this study suggest. Intervention programs fostered more informed and supportive beliefs regarding victim blaming, intervention measures, and trust in victims’ claims of abuse.

Both groups demonstrated positive attitudes toward CSA prevention, with the control group showing higher positivity, compared to the intervention group at the pre-intervention phase). This reflects a general willingness among Nigerian parents to address CSA, consistent with previous findings [22, 27]. Post-intervention, all participants in the intervention group demonstrated positive attitudes. The control group showed a slight, non-significant decrease in scores. These results align with previous studies that highlighted the effectiveness of educational interventions in improving attitudes [28, 29]. Letourneau et al. [30] emphasized the importance of involving parents in CSA prevention programs, as they play a critical role in their children’s sexual development.

In evaluating the self-efficacy of the study participants in CSA prevention, the study shows a significant improvement demonstrated by the participants from the baseline to the 4^th^ and 12^th^ week post-intervention. This agrees with previous studies which reported increased self-efficacy among parents following educational interventions [31, 32]. In a study reported by Campbell [33], 90% of the participants (fathers) talked to their children about CSA prevention, though they might express doubt about their efficacy and competency in having the talk with their children. Participants (fathers) in this study also stated that they believe easily accessible resources will increase their efficacy in talking to their children about CSA prevention, which could lead to the increased protection of children in their environment. The findings from this study indicate that educational interventions can significantly improve the self-efficacy of parents in the prevention of CSA. Importantly, the improved level of self-efficacy post-intervention indicates an increased sense of confidence and empowerment among parents in their ability to recognize, respond to, and prevent instances of CSA within the families and communities. The findings also support Bandura’s self-efficacy theory, which posits that providing individuals with the necessary information and skills enhances their confidence in handling specific situations. Previous studies [5, 34–35] further corroborate these results, emphasizing the role of educational interventions in empowering parents to recognize, respond to, and prevent CSA.

The findings of this study have implications for practice and policy. By prioritizing educational interventions, parents can be equipped with the knowledge and skills required for preventing CSA and promoting the safety and well-being of children. The implication of this study is that educational interventions aimed at promoting CSA intervention should be prioritized and incorporated into curricula and community programs. Policymakers and practitioners should consider the findings of this study when developing programs and policies aimed at preventing CSA.

CSA is primarily the duty of parents and their caregivers. Raising the awareness is the key success factor in avoiding precarious situations, recognizing the signs of CSA, and listening to the child’s problem without skepticism. Parents should take responsibility for their child’s education on social norms and interactions. It is within the home environment the child should learn to reject inappropriate requests and avoid hazardous situations.

## Conclusion

The findings of the study concluded that the intervention was effective in improving the knowledge and attitude of parents toward CSA prevention. It also positively impacted the self-efficacy of parents at CSA prevention, as the intervention has also freely instilled confidence in them to have open communication with their children and perform preventive measures effectively.

The study’s findings support the integration of evidence-based educational interventions into existing healthcare and educational services in Nigeria. In the end, breaking the silence on child sexual abuse in Nigeria is not just about exposing the problem; it is about taking decisive action to end it. It is recommended that educational interventions be widely implemented to raise awareness and empower parents to effectively prevent CSA in their communities.

### Limitations

While this study provides valuable insights into the effectiveness of an educational intervention on parents’ knowledge, attitude, and self-efficacy regarding CSA prevention, several limitations must be acknowledged. First, the quasi-experimental design, though appropriate for this context, lacks random assignment, which limits the ability to make causal inferences. Although random sampling was used in the selection of participants, the non-random assignment of schools to the intervention and control groups could introduce selection bias, potentially affecting the generalizability of the results.

Second, the reliance on self-reported data through questionnaires may be subject to social desirability bias, where participants might over-report positive behaviors or attitudes toward CSA prevention. This limitation is particularly relevant in a cultural context where discussing sexual abuse may still be a taboo subject, influencing the accuracy of responses.

Third, the study was conducted in a specific region of Ogun State, Nigeria, and the findings may not be representative of other regions with different socio-cultural dynamics. The sample was also limited to parents who were able to attend the educational sessions, potentially excluding those who might benefit most, such as parents with lower literacy levels or those unable to attend due to work commitments. Additionally, the study’s follow-up period was only 12 weeks post-intervention, which may not be sufficient to assess the long-term sustainability of the intervention’s effects on parents’ knowledge, attitudes, and self-efficacy. Longer-term follow-up would be needed to determine whether the gains observed are maintained over time and translate into sustained changes in parental behaviours related to CSA prevention.

Lastly, the study did not assess other potentially confounding factors, such as prior exposure to CSA-related education or the socio-economic status of the participants, which could influence the outcomes. Future studies should consider these variables to better understand the broader context in which CSA prevention interventions operate. These limitations suggest that while the educational intervention was effective in the short term, further research using randomized controlled trials, longer follow-up periods, and consideration of additional confounding factors is needed to confirm the long-term effectiveness and applicability of such interventions in diverse contexts.

## Data Availability

Data underlying the results presented in the study will only be made available on request from the author

## Acknowledgments

The author acknowledges the participants and all the staff and management in the study centers; the teachers, principal, vice principals in charge, and research assistants for their voluntary participation, willingness, and cooperation.

